# Study Protocol for a Single-Arm Pilot Feasibility Trial Evaluating a Structured Breathing-Based Intervention to Reduce Burnout and Enhance Mental Well-Being Among Healthcare Professionals in Community-Based Practice Settings

**DOI:** 10.64898/2026.01.07.26343607

**Authors:** Pravesh Sharma, Rowa Osman, Marin Nycklemoe, Danielle M. Boos, Crystal M. Murph, Carolyn Flock, David N. Jacobson, Seokbeen Lim, Jody L. Nation, Melissa A. Wilson, Kristina Schuldt, Paul H. Min

## Abstract

**Introduction:** Burnout among healthcare professionals remains a critical public health issue linked to impaired cognition, emotional exhaustion, and diminished clinical performance. Structured breathing practices have demonstrated promise in improving autonomic regulation and cerebral oxygenation, yet their feasibility and acceptability in real-world healthcare settings remain underexplored.

**Objectives:** This single-arm pilot feasibility trial aims to evaluate the feasibility, acceptability, and implementation appropriateness of a structured breathing-based intervention for healthcare professionals across community-based Mayo Clinic Health System (MCHS) sites. Secondary objectives include assessing usability and engagement with the mobile breathing platform, while exploratory analyses will examine preliminary signals of change in psychological well-being and cerebral hemodynamics.

**Methods and Analysis:** A total of 40 physicians and nurses reporting moderate or greater burnout will be enrolled across four MCHS sites. Participants will complete a four-month structured breathing program delivered primarily online, supported by a mobile application for practice tracking. Assessments will occur at baseline, 2 months, and 4 months, with optional functional near-infrared spectroscopy (fNIRS) measures of cerebral oxygenation collected at baseline and 4 months in a subset. Primary outcomes include (1) recruitment yield, retention, and adherence rates; (2) acceptability and participant satisfaction (survey and qualitative feedback); and (3) implementation appropriateness measured by the Acceptability of Intervention Measure (AIM), Intervention Appropriateness Measure (IAM), and Feasibility of Intervention Measure (FIM). Secondary outcomes assess digital engagement and usability through mobile analytics and the System Usability Scale (SUS). Exploratory outcomes evaluate psychological indicators—burnout, depression, anxiety, perceived stress, sleep, fatigue, professional fulfillment, and resilience—and physiological endpoints (fNIRS). Analyses will be descriptive, summarizing feasibility metrics with 95% confidence intervals. Progression criteria (recruitment ≥75%, retention ≥80%, adherence ≥70%, AIM/IAM/FIM ≥4.0) will determine readiness for a definitive hybrid effectiveness–implementation trial.

**Ethics and Dissemination:** The study is approved by the Mayo Clinic Institutional Review Board (IRB # 25-009320). All participants will provide informed consent. Study procedures ensure confidentiality, cultural sensitivity, and participant safety. Data will be securely stored in REDCap and disseminated through peer-reviewed publications and scientific conference

**ClinicalTrials.gov ID:** The study has been prospectively registered at ClinicalTrials.gov (Identifier NCT07218458).

**Strengths and Limitations:**

- Evaluates a structured breathing-based intervention using both subjective and objective indicators of feasibility, including recruitment, retention, adherence, and implementation appropriateness.
- Integrates mobile application analytics with standardized implementation measures (AIM, IAM, FIM) to capture digital engagement and real-world usability.
- Incorporates functional near-infrared spectroscopy (fNIRS) to explore physiological mechanisms underlying burnout recovery, linking cerebral oxygenation to behavioral outcomes.
- Conducted across diverse community-based Mayo Clinic Health System sites, enhancing ecological validity and relevance to decentralized clinical settings.
- Limitations include the single-arm, region-restricted design, modest sample size, and reliance on volunteer participants, which constrain generalizability and preclude causal inference; exploratory outcomes are intended to guide future trial refinement.

## INTRODUCTION

Occupational distress, including professional burnout has become a growing concern. Burnout refers to an occupational syndrome associated with affective and cognitive changes, including emotional exhaustion, depersonalization or cynicism, and diminished feelings of personal efficacy resulting from chronic occupational stress.^20^ Physician burnout was reported 62.8% in 2021 and remains at nearly 45.2% in 2023, an improvement but still high compared with earlier years.^3, 27^ Burnout in nurses and allied health personnel is likewise prevalent in high-acuity environments. Numerous meta-analyses have estimated the prevalence of nurse burnout, which ranged broadly from 11% to 56%.^35^

Burnout in healthcare professionals is a serious and multifactorial issue influenced by both environmental and individual factors. Increased financial pressures, administrative demands, productivity expectations, workload, and a lack of autonomy are frequently cited as contributing factors.^20^ Furthermore, the rapid advancement of medical knowledge, the implementation of electronic health records, and regulatory requirements create additional pressures. Healthcare professionals also face scrutiny on metrics that may not accurately reflect the complexities of their work.^30^ Burnout among healthcare professionals creates a negative cycle impacting patient care. Consequence includes decreased productivity, impaired connections with patients and colleagues, lower patient satisfaction, and reduced quality of work all contribute to increased work-related stress, further fueling the cycle of burnout.^30^

### Burnout and Chronic Stress Shift Brain Control from Executive Thinking to Emotional Reactivity

The chronic stress associated with factors that are outside the control of an individual (health professionals in this case) exerts detrimental effects on brain circuitry, particularly the prefrontal cortex (PFC). The PFC governs higher-order reasoning, social cognition, and decision-making, including the integration, conceptualization, and critical evaluation of information.^12^ Chronic stress exposure, such as that seen in burnout, causes atrophy of PFC connections, weakening the thoughtful, evaluative responding essential for professional and personal fulfillment.^4, 22^

This impairment in PFC functioning manifests in the workplace as difficulty managing complex tasks, increased potential for medical errors, suboptimal patient care, diminished commitment to professionalism, impaired social connections with patients and colleagues, and a higher likelihood of unprofessional behaviors. ^18, 30^ Sleep deprivation, a common consequence of demanding clinical schedules critically mediates chronic stress–induced PFC dysfunction by exacerbating fatigue and reducing PFC metabolic and physiological activity, thereby contributing to measurable cognitive deficits.^6, 37^

Further, chronic stress can reduce oxygenation of PFC in several ways. The *fight or flight* response triggered by stress causes blood vessels to constrict, reducing blood flow and oxygen delivery to the brain tissues.^33^ Stress leads to shallow, rapid breathing, which decreases the efficiency of oxygen intake.^13^ Functional neuroimaging studies, including functional near-infrared spectroscopy (fNIRS), show that reduced oxygenation in this region correlates with diminished activation during tasks that require inhibitory control, working memory, and complex reasoning.^2, 13^ This hypometabolic state further weakens the PFC’s ability to regulate more primitive brain circuits, such as the amygdala, striatum, and brainstem.^22^ Under these conditions, elevated norepinephrine and dopamine release in these subcortical regions amplifies reflexive habits, emotional reactivity, and threat perception.^4^ This brain state pushes us toward quick, survival-focused reactions, like fear or irritability, instead of calm, deliberate problem solving.^4,22^ While that can be useful in a true emergency, it works against us in the complex, high-stakes situations faced in healthcare. In contrast, when clinicians receive adequate support, stress is more controllable, allowing the PFC to maintain regulatory dominance and foster adaptive coping.

Overall, this neurobiological background highlights the brain’s natural stress response and explains how a diminished sense of control is linked to weakened PFC connections and oxygen metabolism. This pathway forms the mechanistic rationale for targeting PFC oxygenation as an intervention point in burnout.

### Neural Correlates of Breathing Practices

Advanced neuroimaging studies show that breathing rhythms can influence key aspects of brain physiology, including cerebrospinal fluid (CSF) circulation and vasomotor function throughout the brain.^15^ Evidence from electroencephalogram (EEG), functional magnetic resonance imaging (fMRI), and fNIRS supports that structured breathing can modulate neural oscillations, cerebral perfusion, and autonomic tone, all of which have direct implications for PFC health and executive control.^38^

### Enhanced Brain Oxygenation and Modulation of Brain Activity

Controlled and structured breathing patterns have been shown to increase cerebral oxygen levels and stabilize brain activity, contributing to improved emotional regulation and cognitive performance.^9–10, 17^ One important physiological marker of this effect is heart rate variability (HRV)—the natural variation in time between heartbeats—which reflects parasympathetic nervous system activity and is closely linked to stress regulation and resilience.^29^

Structured breathing has been found to increase alpha wave activity (8–13 Hz), which is associated with relaxed, focused attention, and to decrease theta wave activity (4–8 Hz), which is linked to mental noise and rumination.^38^ Functional MRI studies demonstrate that structured breathing enhances the BOLD (Blood-Oxygen-Level Dependent) signal in cortical and subcortical areas such as the medulla and hippocampus, regions where activation is positively correlated with HRV.^11,29^ In contrast, HRV is negatively correlated with activity in the anterior insula, dorsomedial prefrontal cortex, and occipital cortex.

These neural and physiological changes are directly relevant to burnout, where chronic stress and sleep deprivation weaken PFC connections and reduce its ability to regulate primitive brain circuits such as the amygdala, striatum, and brainstem.^4^ By improving cerebral oxygenation and strengthening functional connectivity within the PFC, structured breathing may help restore executive regulation and reduce maladaptive, reactive brain states (Figure 1).^9–10^

**Figure 1.**
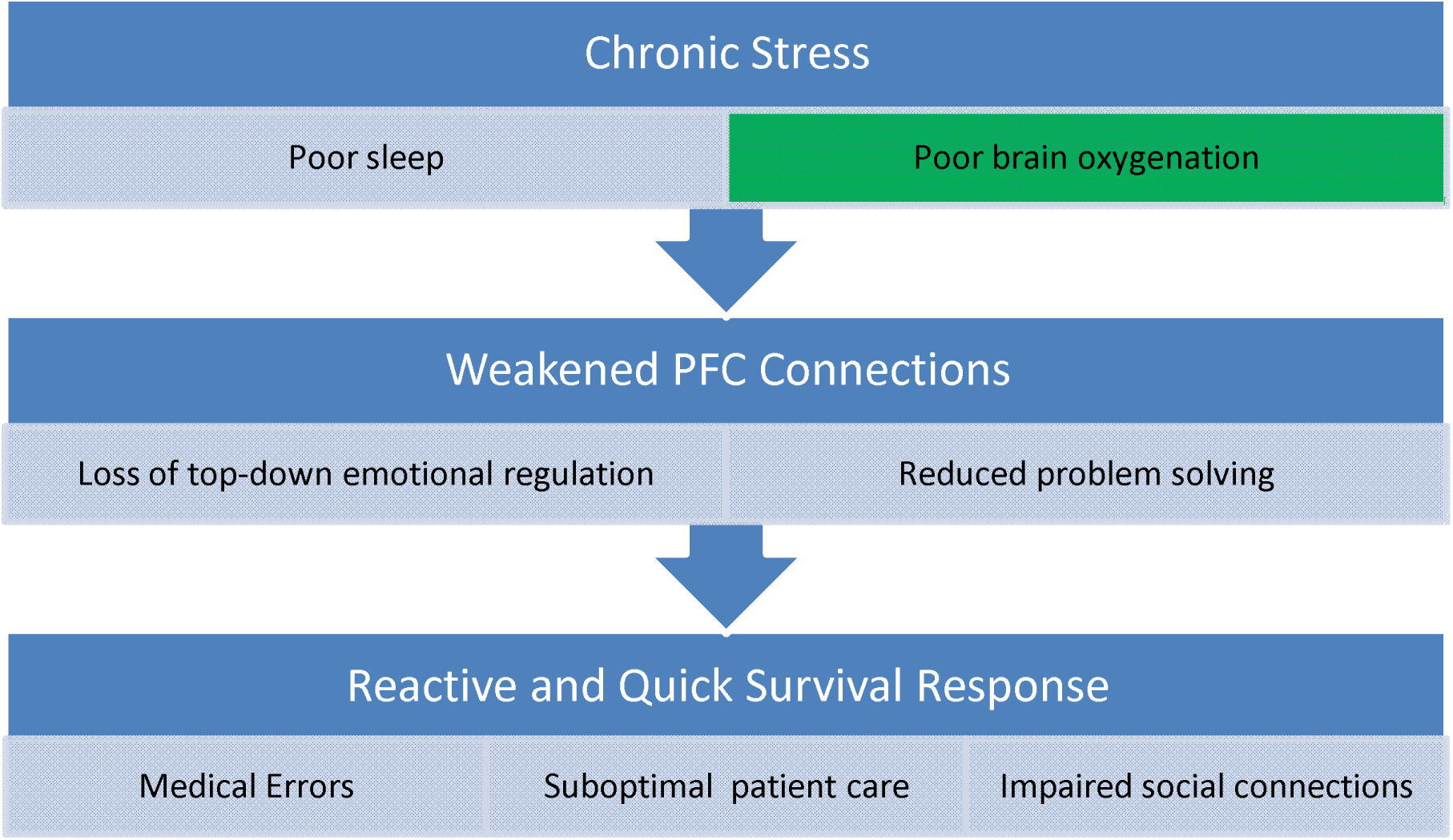
Proposed neurophysiological pathway linking burnout to impaired executive control and heightened emotional reactivity. Chronic stress and sleep deprivation reduce prefrontal cortex (PFC) oxygenation, weakening its regulatory control over primitive brain circuits (amygdala, striatum, brainstem). This shift favors rapid, emotionally driven responses over deliberate reasoning. Color codes, GREEN box is intervention target. The intervention focuses on improving PFC oxygenation, which may not only restore executive regulation but could also enhance sleep quality as a downstream benefit.

### Cerebrospinal Fluid Dynamics During Breathing Practice

CSF and interstitial fluid (ISF) are needed to maintain central nervous system (CNS) homeostasis. Beyond its mechanical cushioning role, CSF also facilitates the distribution of nutrients and hormones within the CNS. CSF and ISF collectively play a critical role in removing solutes and metabolic waste from interstitial spaces.^11^ This is an important function implicated in the pathophysiology of a vast array of neurodegenerative and neuroinflammatory conditions.

CSF flow is controlled primarily by alterations in the CNS vascular bed, including cardiac pulsations and respiration, as well as secondarily by alterations in body position and the cough reflex.^16^ Deep inspiration creates a forceful upward flow of CSF from the lumbar space up the spinal canal into the cranial vault. With expiration, CSF flows partially downward, creating a rhythmic bidirectional flow. The fall in intrathoracic pressure on inspiration enables venous drainage from the brain and neck into the thoracic cavity. This temporary reduction of intracranial blood volume is compensated by an upward shift of CSF to maintain intracranial pressure equilibrium, as per the *Monro–Kellie doctrine*. Deep, structured breathing thus produces dynamic pressure changes that actively drive CSF circulation—upward on inspiration and downward on expiration.^36^ By enhancing CSF circulation, structured breathing may improve oxygen and nutrient delivery to the PFC while aiding in the clearance of metabolic byproducts, supporting recovery from burnout-related neural fatigue. ^14–15, 36^

While breathing training is rapidly gaining attention as a potential intervention to boost brain function and autonomic regulation, few studies have integrated objective diaphragm movement assessment, fNIRS measurement of cortical oxygenation, and validated mental health/burnout scales in the same protocol.^19–10, 25, 28^ Our method bridges this gap and aims to establish a clinically applicable breathing practice program for reducing and preventing burnout in healthcare professionals.

#### Research Question

Can a structured breathing-based intervention be implemented and accepted among healthcare professionals, as reflected by satisfactory recruitment, retention, adherence, and participant satisfaction? Additionally, how usable and engaging is the mobile application platform for supporting intervention delivery and monitoring? Exploratorily, what are the preliminary physiological and mental health effects of the breathing-based intervention, including changes in cerebral oxygenation and local tissue hemodynamics measured by fNIRS, to inform the design of a future larger-scale trial?

### Study Objectives

#### Primary Objective

To assess the feasibility, participant satisfaction, acceptability, and implementation appropriateness of a structured breathing-based intervention among healthcare professionals, including physicians and nurses. Feasibility will be determined by the ease of recruitment, and retention through the final assessment. Participant satisfaction will be assessed through a mixed-methods approach combining quantitative surveys and qualitative participant interviews.

Acceptability and implementation appropriateness will be evaluated using satisfaction ratings and standardized implementation outcome measures, including the Acceptability of Intervention Measure (AIM), Intervention Appropriateness Measure (IAM), and Feasibility of Intervention Measure (FIM).

#### Secondary Objective

To evaluate intervention adherence, usability, and engagement metrics collected through the mobile application platform. These data will be used to identify barriers and facilitators to digital engagement, assess usability, and inform refinements to the intervention’s delivery and technological infrastructure for future large-scale trials.

#### Exploratory Objective

To assess the physiological effects of the breathing-based intervention on cerebral oxygenation and local tissue hemodynamics, as measured by functional near-infrared spectroscopy (fNIRS), along with changes in mental health outcomes. These exploratory analyses are intended to assess the feasibility of collecting physiological and psychological data, examine preliminary trends, and estimate variability and effect sizes that can inform the design and power calculations of future definitive trials. <u>The study is not powered to test effectiveness or efficacy hypotheses at this stage.</u>

## METHODS

This protocol adheres to the Standard Protocol Items: Recommendations for Interventional Trials (SPIRIT) 2013 statement for the reporting of clinical trial protocols.^5, 32^ Relevant guidance from the CONSORT 2010 Extension for Pilot and Feasibility Trials was also integrated to align with the feasibility objectives and implementation-focused nature of this study.^8^

As a pilot feasibility study, this trial is not designed to test hypotheses but to evaluate whether the structured breathing-based intervention can be implemented effectively within a healthcare setting.^23^ Progression to a future large-scale, hybrid effectiveness–implementation trial will be based on meeting *predefined feasibility benchmarks*. These include recruitment of at least 75% of eligible participants, retention of 80% or more through the 4-month follow-up, mean Acceptability of Intervention Measure (AIM), Intervention Appropriateness Measure (IAM), and Feasibility of Intervention Measure (FIM) scores of 4.0 or higher on a 5-point scale, and adherence of at least 70% to self-reported or app-logged breathing sessions.^21, 31, 34^ Achieving these thresholds will indicate readiness to scale the intervention, while values within approximately 10 percentage points of these targets will signal the need for protocol modification before proceeding to a definitive trial.

### Study Design

This is a single-arm, non-randomized pilot feasibility study designed to evaluate the feasibility, participant satisfaction, acceptability, and implementation appropriateness (primary objective), as well as engagement (secondary objective) and preliminary efficacy signals (exploratory objective) of a structured breathing-based intervention on physiological and psychological indicators of burnout among healthcare professionals, including physicians and nurses.

### Study Setting

This study is planned to be conducted at the Mayo Clinic’s community-based care setting provided by Mayo Clinic Health System (MCHS), which serves patients across urban and rural settings primarily in the Upper Midwest.^1^ The MCHS has four large hospitals, two each in MN and Wisconsin where this study will be conducted. MCHS service area presents with rural health challenges as characterized by high disease burden, and patient flow, limited specialty excess and growing need for health innovations. The MCHS is staffed by >1,000 physicians and >14,000 allied staff across MN, WI and IA. MCHS includes 16 hospitals (including 10 rural critical access hospitals), 41 multispecialty clinics, and a mobile health clinic, and supports 2 million patient visits annually.

### Participants and Recruitment

#### Inclusion criteria

This single-arm intervention trial will enroll healthcare professionals who meet the following eligibility criteria: 1) Currently employed as a physician or nurse at one of the four designated MCHS sites: Eau Claire, WI; La Crosse, WI; Mankato, MN; or Albert Lea, MN, 2) score greater than 40 on the Copenhagen Burnout Inventory (questions 1–6), 3) physically able to perform light exercise, and 4) have access to a smartphone or tablet. We plan to recruit N = 40 healthcare professionals, targeting an equal distribution of physicians and nurses (1:1 ratio when possible).^16^

#### Exclusion Criteria

The exclusion criteria include: 1) severe or unstable medical condition that could interfere with participation or data collection, 2) active neurological condition (including seizure disorder, traumatic brain injury, or stroke) that could affect cognitive functioning or brain imaging results, 3) chronic lung disease (e.g., chronic obstructive pulmonary disease (COPD), cystic fibrosis) or aneurysm, and 4) current pregnancy or planning to become pregnant during the study period.

Participants are asked to avoid caffeinated products on the morning of the fNIRS study visits. Stay hydrated but avoid excessive drinking. It would be fine to have a light meal before the experiment but avoid overeating. To accommodate potential scheduling conflicts, there will be a permissible leeway of up to three weeks for each visit. Due to potential scheduling conflicts with participants’ work schedules, we will allow a 3-week leeway for completing visits.

### Outreach and Engagement Plan

To facilitate recruitment and increase awareness of the study, we will host a Breathing Symposium at each of the four participating MCHS sites—Eau Claire, WI; La Crosse, WI; Mankato, MN; and Albert Lea, MN. These symposia will be open to all clinicians and healthcare staff and will focus on:

- Educating attendees about burnout, its neurobiological basis, and the impact of chronic stress on brain health.
- Presenting the scientific evidence for structured breathing techniques as a potential intervention for improving mental well-being.
- Demonstrating breathing practices and their physiological effects.
- Providing a live fNIRS demonstration to show how brain oxygenation and deoxygenation can be measured in real time during breathing activities.

### On-Site Recruitment Activities

A study-specific Clinical Research Coordinator (CRC) will be present at each symposium to:

1. Provide printed flyers with study details, eligibility criteria, and contact information for the research team.
2. Engage with interested attendees, answer questions, and offer initial eligibility screening information.
3. Collect contact details from potential participants who express interest in enrolling.

The MCHS Well-Being Office will distribute an email announcement to all MCHS employees when recruitment begins. These combined in-person and electronic outreach strategies are designed to maximize awareness, ensure broad reach across the participating sites, and facilitate balanced enrollment of physicians and nurses.

### Intervention

Each participant will undergo a four month online systematic breathing intervention and complete the pre- and post-intervention surveys, and fNIRS. No randomization or allocation concealment is used, as this is a single arm pilot feasibility study. CRC and principal investigator (PI) will review attendance logs and app analytics monthly to ensure the intervention is delivered as planned.

### Breathing Practice Protocol

The structured online and in-person breathing practices (in-person practice is not required for this study) will be led by experienced instructors. The breathing practice focuses on conducting soft and gentle exercising, to help participants naturally relax and strengthen the core muscles necessary for better breathing control, thereby improving one’s everyday natural breathing pattern over time. It is designed to be suitable for all participants, ensuring accessibility and ease of practice. The program supports online video modules so participants can follow and practice at their own pace, supplemented by standardized written materials that outline the practice principles and postures. The breathing practice applied in the study is based on a beginner level program. Adopting a structured breathing practice program helps avoid potential biases from experimenter-led sessions, ensuring consistency and reproducibility.

Participants will register and enroll in the official breathing program, joining their regular practice sessions like any other members, without disclosing their participation in the study to fellow members. The breathing practice instructors will be informed about the participants’ enrollment in the study, specifically their first name and age, but will not be given any other study details. No direct identifiers will be shared. No post-trial intervention is provided; participants receive standard community referrals and treatment after the completion of this trial.

### Breathing Practice Sessions for Participants

- (Required) Participants will complete two real-time online sessions with breathing instructors and the study team: an initial instruction session and a follow-up session, each about 60 minutes. The follow-up session should take place within 45 days of the first session. Depending on availability, the initial instruction session may be conducted in person.
- (Required) Participants are encouraged to engage in 15–36-minute self-practice sessions, ideally three times per week, with a minimum of one session per week. During the initial session, a stepwise approach will be discussed with participants to help set realistic goals, starting with an easy and manageable schedule and building up gradually as they feel comfortable.
- (Optional) Weekly real-time online practice sessions, conducted via video conference, are available to provide live, interactive instruction in the breathing practices. These sessions complement self-practice by offering real-time guidance, progress evaluation, and individualized feedback from instructors. More than 10 weekly sessions are offered. Feedback will be shared with both participants and the study team to monitor training effectiveness and adaptation. Participants are encouraged to continue attending these weekly sessions as an option.

Participants are encouraged to regularly communicate with instructors and the study team about their progress and any questions they may have about the practice. The study team will respond to communications and prompt participants appropriately. This routine serves as a check-in to encourage regular engagement with the breathing exercises and to help maintain a record of participation. Participants may also increase the frequency of their daily practice sessions if desired.

### Data Collection Methods

The data will be collected through fNIRS recording app and, RedCap surveys (noted in outcomes sections) and mobile application.

### fNIRS Assessment Methods

fNIRS will be used to measure cerebral hemodynamic patterns in participants, focusing on changes in brain activity in the prefrontal cortex, which is involved in mood regulation and cognitive function.

Detailed information about the fNIRS device (Obelab, Inc.) and specific task protocols is provided in the Device Information and Procedures sections of the protocol (Figure 2).^24^ The brain imaging acquisition will take about 30 minutes to complete, including placing the device over the head and completing resting-state and task-based measurements. The task-based measurement includes word fluency test and deep breathing session.

**Figure 2.**
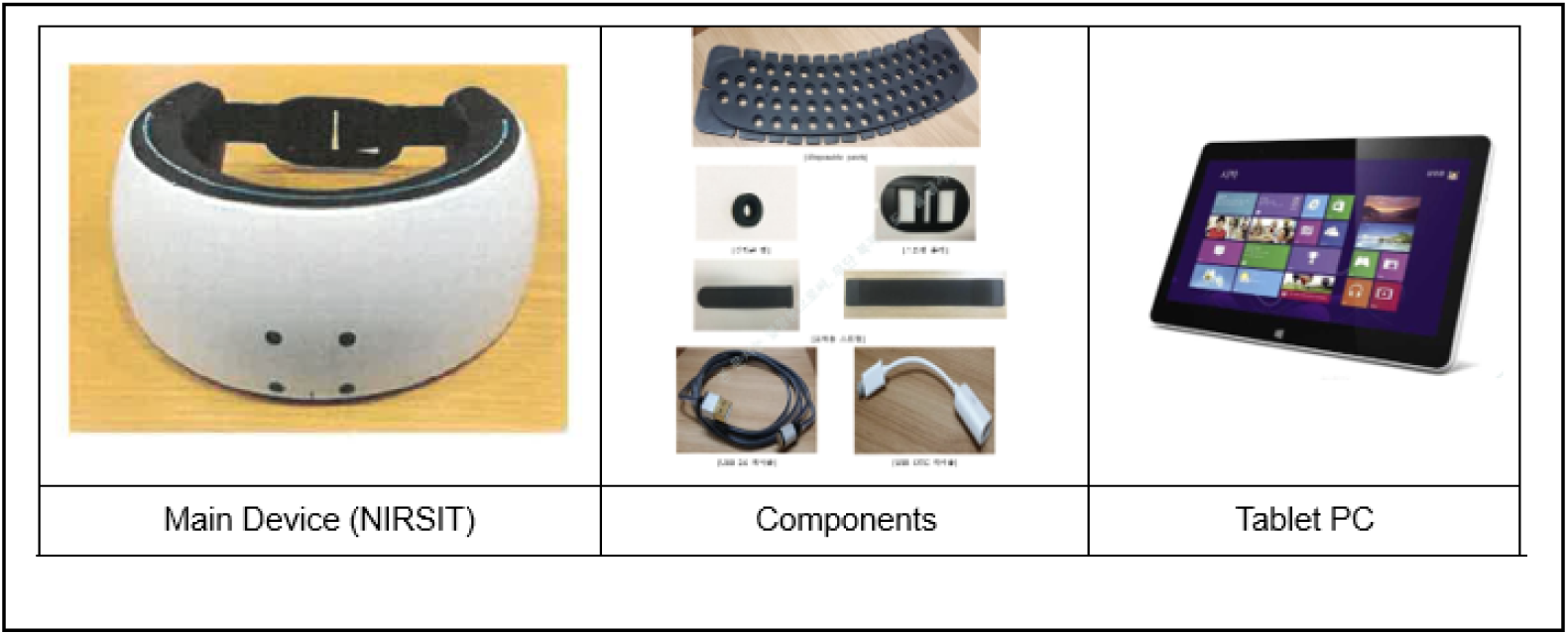
functional near-infrared spectroscopy (fNIRS) device and tablet. fNIRS uses infrared technology to measure oxyhemoglobin levels in the circulation, this enables the device to measure cerebral hemodynamic patterns in real time.

During the study, the subject wears the fNIRS device, and the researcher runs the software on the tablet screen to measure changes in the subject’s oxyhemoglobin.

#### Adherence and Compliance

Mobile breathing app will track adherence and compliance. During one of the in-person meetups, we will go through how to use the mobile app and provide instruction.

1. Participant Engagement: Participants will receive biweekly breathing tips through email, offering encouragement and an opportunity to ask any questions. If a participant does not communicate with the study team for 5 days on their daily practice, the study team can send a reminder via their preferred contact method (i.e., text message, telephone call, or email). This ensures ongoing engagement and addresses any participant concerns. For the breathing group, any feedback from the instructors will be relayed to the participants, and vice versa.
2. Emphasis on Honest Reporting: We encourage participants to report their practice habits honestly and without any pressure. Self-practice is not mandatory, and there’s no need to overcommit. What’s most important is that participants provide genuine feedback on their actual practice. The study seeks to understand natural compliance patterns, and we value honest, pressure-free reporting over strict adherence to practice frequency.

### Data collection timeline

This project allows participants to complete their visit related activities online or in person (Table 1).

**Table 1.** Schedule of Enrollment, Interventions, and Assessments (Participant Timeline)

| **TRIAL PERIOD** | **Enrollment** | **Post-Enrollment** | **Follow-up** | **Close-out** |
| --- | --- | --- | --- | --- |
| **TIMEPOINT** | **-t<sub>0</sub> to 0** | **0 months (Visit 1)** | **2 months (Visit 2)** | **4 months (Visit 3)** |
| **ENROLLMENT:** |  |  |  |  |
| Eligibility screen | X |  |  |  |
| Informed consent | X |  |  |  |
| **INTERVENTION:** |  |  |  |  |
| Mobile breathing-based intervention (setup, registration, practice) |  | X |  |  |
| Continued app-guided breathing practice |  | → | → | → |
| **ASSESSMENTS:** |  |  |  |  |
| Survey questionnaires (REDCap) |  | X | X | X |
| fNIRS (functional near-infrared spectroscopy; site-specific) |  | X |  | X (optional) |
| Participant satisfaction and usability (embedded in surveys) |  | X | X | X |
| Adherence monitoring via mobile app |  | → | → | → |

### Outcomes

The primary outcomes of this feasibility study focus on implementation-related domains, including (1) recruitment and retention metrics derived from study logs (e.g., accrual velocity, time to first enrollment, and completion rates); (2) acceptability and participant satisfaction, assessed through structured surveys and qualitative interviews at the end of the 4-month intervention; and (3) implementation appropriateness and feasibility of intervention delivery, measured using the Acceptability of Intervention Measure (AIM), Intervention Appropriateness Measure (IAM), and Feasibility of Intervention Measure (FIM) at 2 and 4 months.

The secondary outcome pertains to usability and engagement with the mobile platform, assessed through mobile application analytics (logins, session duration, and completion rates) and the System Usability Scale (SUS). These data will be captured continuously through automated tracking and summarized at 4 months.

Exploratory outcomes include changes in psychological and physiological indicators from baseline to 2- and 4-month follow-up. Psychological indicators encompass burnout (Copenhagen Burnout Inventory), mental health (Patient Health Questionnaire-9, Generalized Anxiety Disorder-7, Perceived Stress Scale), sleep and fatigue (Pittsburgh Sleep Quality Index, Modified Fatigue Impact Scale), and professional well-being (Professional Fulfillment Index, Linear Analog Self-Assessment, Brief Resilience Scale). Physiological outcomes will be derived from functional near-infrared spectroscopy (fNIRS) assessments of cerebral oxygenation and hemodynamics, obtained at baseline and 4 months in a subset of participants.

All outcomes and corresponding assessment schedules are summarized in table 2.

**Table 2.**
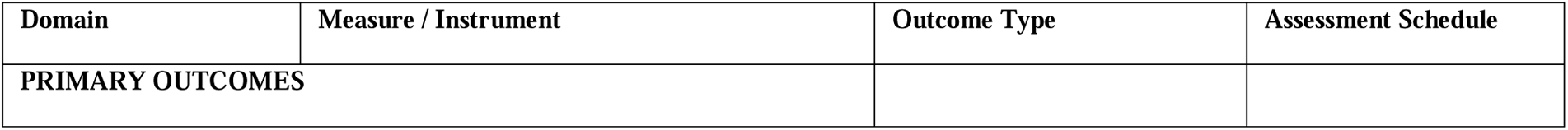
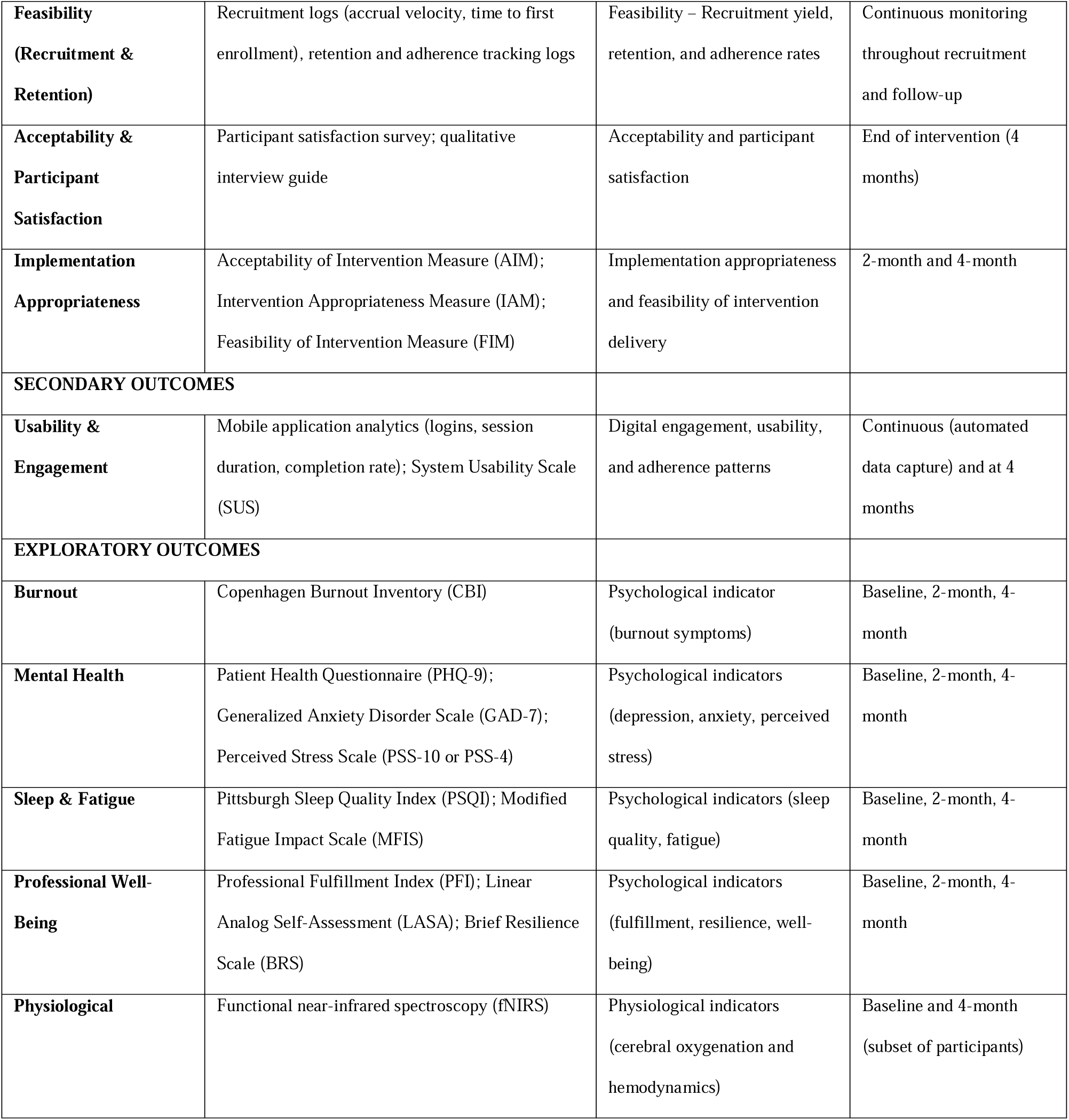
Summary of Study Measures, Instruments, and Assessment Schedule.

| Domain | Measure / Instrument | Outcome Type | Assessment Schedule |
| --- | --- | --- | --- |
| **PRIMARY OUTCOMES** |  |  |  |
| **Feasibility**<br>**(Recruitment & Retention)** | Recruitment logs (accrual velocity, time to first enrollment), retention and adherence tracking logs | Feasibility – Recruitment yield, retention, and adherence rates | Continuous monitoring throughout recruitment and follow-up |
| **Acceptability & Participant Satisfaction** | Participant satisfaction survey; qualitative interview guide | Acceptability and participant satisfaction | End of intervention (4 months) |
| **Implementation Appropriateness** | Acceptability of Intervention Measure (AIM);<br>Intervention Appropriateness Measure (IAM);<br>Feasibility of Intervention Measure (FIM) | Implementation appropriateness and feasibility of intervention delivery | 2-month and 4-month |
| **SECONDARY OUTCOMES** |  |  |  |
| **Usability & Engagement** | Mobile application analytics (logins, session duration, completion rate); System Usability Scale (SUS) | Digital engagement, usability, and adherence patterns | Continuous (automated data capture) and at 4 months |
| **EXPLORATORY OUTCOMES** |  |  |  |
| **Burnout** | Copenhagen Burnout Inventory (CBI) | Psychological indicator (burnout symptoms) | Baseline, 2-month, 4-month |
| **Mental Health** | Patient Health Questionnaire (PHQ-9);<br>Generalized Anxiety Disorder Scale (GAD-7);<br>Perceived Stress Scale (PSS-10 or PSS-4) | Psychological indicators (depression, anxiety, perceived stress) | Baseline, 2-month, 4-month |
| **Sleep & Fatigue** | Pittsburgh Sleep Quality Index (PSQI); Modified Fatigue Impact Scale (MFIS) | Psychological indicators (sleep quality, fatigue) | Baseline, 2-month, 4-month |
| **Professional Well-Being** | Professional Fulfillment Index (PFI); Linear Analog Self-Assessment (LASA); Brief Resilience Scale (BRS) | Psychological indicators (fulfillment, resilience, well-being) | Baseline, 2-month, 4-month |
| **Physiological** | Functional near-infrared spectroscopy (fNIRS) | Physiological indicators (cerebral oxygenation and hemodynamics) | Baseline and 4-month (subset of participants) |

### Participant Retention, Withdrawal, and Discontinuation

Participant retention will be promoted through flexible scheduling, regular reminders from the CRC, and engagement messages through the study’s mobile application. Any participant who chooses to withdraw from the study or discontinues the intervention will be asked, if willing, to complete a brief exit survey to document their reasons for withdrawal (e.g., time constraints, technical issues, loss of interest, or adverse experience). Data collected up to the point of withdrawal—including baseline and interim assessments—will be retained and included in descriptive analyses in accordance with SPIRIT 2013 recommendations. The CRC will record all withdrawals in the study tracking log, and the PI will review these monthly to identify potential barriers to retention. No replacement participants will be enrolled once recruitment is complete.

### Statistical Analysis

Analyses will follow the CONSORT 2010 Extension for Pilot and Feasibility Trials and the SPIRIT 2013 guidelines, emphasizing feasibility, acceptability, and implementation readiness rather than hypothesis testing. Because this is a pilot feasibility study, formal statistical hypothesis testing and power calculations are not planned. Instead, all quantitative outcomes will be summarized descriptively using means, standard deviations, medians with interquartile ranges, and proportions with 95% confidence intervals, as appropriate. Progression criteria, rather than p-values, will guide interpretation of feasibility and readiness for scale-up, consistent with the *National Center for Complementary and Integrative Health (NCCIH)* guidance on pilot studies.^23^ Meeting the prespecified benchmarks for recruitment, retention, adherence, and acceptability will indicate that the intervention is feasible and warrants evaluation in a fully powered hybrid effectiveness–implementation trial.

All analyses will be conducted using standard statistical software (e.g., R). Continuous variables will be summarized as means with standard deviations or medians with interquartile ranges, and categorical variables as frequencies and percentages. Feasibility will be evaluated based on prespecified thresholds: recruitment of at least 75% of eligible participants, retention of ≥80% through the 4-month follow-up, and adherence of ≥70% to self-reported or app-logged breathing sessions. Recruitment rate (participants per week), time to first enrollment, retention, and adherence will be summarized descriptively with 95% confidence intervals. Meeting or exceeding these thresholds will indicate feasibility of scaling the intervention. Acceptability and implementation appropriateness will be assessed using mean scores from the Acceptability of Intervention Measure (AIM), Intervention Appropriateness Measure (IAM), and Feasibility of Intervention Measure (FIM), complemented by qualitative feedback analyzed thematically to identify barriers and facilitators.

Mobile-application usability and engagement metrics (e.g., logins, session duration, completion rate) will be summarized descriptively. Exploratory outcomes—including burnout, depression, anxiety, perceived stress, sleep, fatigue, well-being, and physiological measures from fNIRS—will be analyzed descriptively across baseline, 2-month, and 4-month assessments. Changes over time will be reported as mean or median differences with 95% confidence intervals to estimate variability and potential trends. No formal hypothesis testing will be conducted. Findings will be interpreted descriptively, and estimates will be used to guide effect-size estimation, power calculation, and protocol refinement for a future fully powered hybrid effectiveness–implementation trial.

## Results

The results of this feasibility project will establish whether a structured breathing-based intervention can be implemented efficiently and accepted by healthcare professionals in a busy clinical environment. Findings will inform the scalability of the intervention as a practical tool for addressing burnout and supporting workforce well-being across the health system. Data generated from this pilot will provide preliminary parameters—such as recruitment yield, adherence rates, retention, and participant satisfaction—that will guide the design, implementation logistics, and power estimation of future large-scale randomized or comparative effectiveness trials.

## DISCUSSION

### Principal Findings

This single-arm pilot feasibility study will evaluate the practicality, acceptability, and preliminary signals of impact of a structured breathing-based wellness intervention for healthcare professionals. Feasibility will be assessed through prespecified quantitative thresholds for recruitment, retention, adherence, and acceptability, complemented by qualitative feedback to understand barriers and facilitators of participation. Exploratory analyses will describe trends in burnout, stress, and well-being indices. These feasibility metrics are consistent with established criteria used in behavioral, digital health, and implementation-focused pilot trials and will inform key design parameters for a subsequent randomized study.

The inclusion of fNIRS represents a novel methodological innovation. fNIRS enables the quantification of cortical oxygenation and hemodynamic responses associated with the breathing intervention, offering an objective physiological correlate of engagement and mental recovery. Incorporating this neurophysiological measure provides an opportunity to explore early mechanistic signals and enhance participant motivation through awareness of tangible biological effects. To our knowledge, no previous feasibility study has integrated fNIRS as both an implementation and exploratory outcome measure in a wellness intervention targeting clinician burnout.

## Conclusions

This feasibility study will generate foundational evidence on the acceptability, adherence, and implementation appropriateness of a structured breathing-based intervention for healthcare professionals. It will also provide initial data linking behavioral and neurophysiological outcomes through fNIRS assessment. The findings will directly inform power estimation, procedural refinements, and endpoint selection for a future randomized controlled trial aimed at testing both mechanistic and clinical impacts on burnout, resilience, and workplace well-being.

### Trial Oversight and Monitoring

Given the minimal-risk, behavioral nature of this pilot feasibility study, no independent Data Monitoring Committee (DMC) will be established. Study safety, data integrity, and ethical conduct will be overseen by the Principal Investigator and the Mayo Clinic Institutional Review Board (IRB). Oversight procedures will include monthly reviews by the CRC to ensure data completeness and participant safety, as well as quarterly evaluations by the Principal Investigator to monitor protocol adherence, recruitment progress, and any adverse event reports. All protocol amendments will be promptly communicated to the Mayo Clinic IRB, ClinicalTrials.gov, and all study personnel before implementation. Participants will provide written informed consent for survey, interview, and physiological data collection. No biospecimens or genetic testing will be conducted. Confidentiality will be safeguarded through the use of secure, password-protected RedCap databases, and all data will be de-identified prior to analysis and reporting.

### Ethics and Dissemination

This study has been reviewed and approved by the Mayo Clinic Institutional Review Board (IRB #25-009320). The trial will be registered on ClinicalTrials.gov prior to participant enrollment. Participants will receive $50 for each completed in-person visit (Baseline at 0 months and follow-up at 4 months) and $25 for the online visit (2 months). The maximum total remuneration for completing all study visits is $125. Payments will be provided after each visit is completed. All participants will provide informed consent prior to participation, and study procedures have been designed to minimize risk and protect confidentiality. Participant safety will be safeguarded by trained CRCs. Given the behavioral and minimal-risk nature of this feasibility study, serious adverse events (SAEs) are not anticipated. However, any untoward medical or psychological occurrence temporally associated with study participation—such as dizziness, light-headedness, hyperventilation, emotional distress, or unexpected discomfort—will be considered an adverse event (AE). The CR) will document all AEs in the study log, noting onset, duration, resolution, and relationship to the intervention. Participants experiencing significant distress during a session will be offered a brief debriefing and referred to appropriate support resources as outlined in the protocol safety plan. The PI will review AE logs monthly, and all SAEs or unexpected events will be reported promptly to the Mayo Clinic IRB in accordance with institutional policy. Aggregate AE data will be included in study progress reports and final dissemination materials. All data will be stored securely in REDCap and accessible only to authorized study personnel. Data management will adhere to Mayo Clinic policies and NIH data-sharing requirements.

Study findings will be disseminated through peer-reviewed publications, conference presentations, and community reports shared with participants, and local stakeholders. A plain-language summary of results will also be posted on ClinicalTrials.gov. De-identified datasets may be deposited in a suitable research repository in accordance with Mayo Clinic policy and funder requirements.

## Author’s contribution

PS and PHM contributed to funding acquisition, conceptualization, methodology, resources, and writing (original draft, review, and editing). PHM additionally contributed to providing resources, including access to the functional near-infrared spectroscopy (fNIRS) system. DMB, CMM, and CM contributed to project administration, research coordination, and writing (review and editing). RO, DNJ, SL, JLN, MAW, and KS contributed to manuscript drafting, review, and editing. All authors reviewed and approved the final manuscript and agree to be accountable for all aspects of the work.

## Data Availability

All data produced in the rpesent study are available upon reasonable request to the authors

## REFERENCES

1. About Mayo Clinic Health System. Mayo Clinic Health System. Accessed November 11, 2025. https://www.mayoclinichealthsystem.org/about-us

2. Al-Shargie F, Katmah R, Tariq U, Babiloni F, Al-Mughairbi F, Al-Nashash H. Stress management using fNIRS and binaural beats stimulation. Biomed Optics Ex. 2022;13(6):3552–3575. doi:10.1364/BOE.455097

3. American Medical Association. Measuring and addressing physician burnout. AMA. Updated May 15, 2025. Accessed September 20, 2025. https://www.ama-assn.org/practice-management/physician-health/measuring-and-addressing-physician-burnout.

4. Arnsten AF. Stress signaling pathways that impair prefrontal cortex structure and function. Nat Rev Neurosci. 2009;10(6):410–422. doi:10.1038/nrn2648

5. Chan AW, Tetzlaff JM, Altman DG, et. Al. SPIRIT 2013 statement: defining standard protocol items for clinical trials. Ann Intern Med. 2013;158(3):200–7. doi: 10.7326/0003-4819-158-3-201302050-00583.

6. Cribbet MR, Thayer JF, Jarczok MN, Fischer JE. High-Frequency Heart Rate Variability Is Prospectively Associated With Sleep Complaints in a Healthy Working Cohort. Psychoso Med. 2024;86(4):342–348. doi:10.1097/PSY.0000000000001302

7. Deligkaris P, Panagopoulou E, Montgomery AJ, Masoura E. Job burnout and cognitive functioning: A systematic review. Work & Stress. 2014;28(2):107–123.

8. Eldridge S M, Chan C L, Campbell M J, Bond C M, Hopewell S, Thabane L, et. Al. CONSORT 2010 statement: extension to randomised pilot and feasibility trials. BMJ. 2016; 355:i5239 doi:10.1136/bmj.i5239

9. Fincham GW, Kartar A, Uthaug MV, et al. High ventilation breathwork practices: An overview of their effects, mechanisms, and considerations for clinical applications. Neurosci Biobehavi Review. 2023;155:105453. doi:10.1016/j.neubiorev.2023.105453

10. Fincham GW, Strauss C, Montero-Marin J, Cavanagh K. Effect of breathwork on stress and mental health: A meta-analysis of randomized-controlled trials. Scientific Reports. 2023;13(1):432. doi:10.1038/s41598-022-27247-y

11. Gerritsen RJS, Band GPH. Breath of Life: The Respiratory Vagal Stimulation Model of Contemplative Activity. Front Human Neurosci. 2018;12:397. doi:10.3389/fnhum.2018.00397

12. Hölzel BK, Lazar SW, Gard T, Schuman-Olivier Z, Vago DR, Ott U. How Does Mindfulness Meditation Work? Proposing Mechanisms of Action From a Conceptual and Neural Perspective. Persp Psychol Sci. 2011;6(6):537–559. doi:10.1177/1745691611419671

13. Homma I, Masaoka Y. Breathing rhythms and emotions. Experi Physi. 2008;93(9):1011–1021. doi:10.1113/expphysiol.2008.042424

14. Iliff JJ, Wang M, Liao Y, et al. A paravascular pathway facilitates CSF flow through the brain parenchyma and the clearance of interstitial solutes, including amyloid β. Science Translational Medicines. 2012;4(147):147ra111. doi:10.1126/scitranslmed.3003748

15. Kandimalla M, Shmuel A. Cardiorespiratory dynamics in the brain: Review on the significance of cardiovascular and respiratory correlates in functional MRI signal. Neuroimage. 2025;306:121000.

16. Kiviniemi V, Wang X, Korhonen V, et al. Ultra-fast magnetic resonance encephalography of physiological brain activity - Glymphatic pulsation mechanisms?. J Cere Flow Metab. 2016;36(6):1033–1045. doi:10.1177/0271678X15622047

17. Korkmaz A, Bernhardsen GP, Cirit B, et al. Sudarshan Kriya Yoga Breathing and a Meditation Program for Burnout Among Physicians: A Randomized Clinical Trial. JAMA. 2024;7(1):e2353978. doi:10.1001/jamanetworkopen.2023.53978

18. Koutsimani P, Montgomery A, Masoura E, Panagopoulou E. Burnout and Cognitive Performance. International Journal of Environ Res Pub Health. 2021;18(4):2145. doi:10.3390/ijerph18042145

19. Kristensen, T. S., Borritz, M., Villadsen, E., & Christensen, K. B. The Copenhagen Burnout Inventory: A new tool for the assessment of burnout. Work & Stress. 2005;19(3):192–207. 10.1080/02678370500297720

20. Maslach, C., & Leiter, M. P. The truth about burnout: How organizations cause personal stress and what to do about it. Jossey-Bass/Wiley; 1997.

21. Mbuagbaw L, Chen LH, Aluko E, et. Al. Empirical progression criteria thresholds for feasibility outcomes in HIV clinical trials: a methodological study. Pilot Feasibility Stud. 2023;9(1):96. doi: 10.1186/s40814-023-01342-x.

22. McEwen BS, Morrison JH. The brain on stress: vulnerability and plasticity of the prefrontal cortex over the life course. Neuron. 2013;79(1):16–29. doi:10.1016/j.neuron.2013.06.028

23. National Center for Complementary and Integrative Health. Pilot Studies: Common Use and Missuses. NCCIH.NIH. Accessed November 10, 2025. https://www.nccih.nih.gov/grants/pilot-studies-common-uses-and-misuses#:~:text=The%20goal%20of%20pilot%20studies,to%20reasonably%20evaluate%20feasibility%20goals

24. Obelab. Obelab.com. Accessed November 20, 2025. https://www.obelab.com/

25. Park YJ. Association of autonomic function and brain activity with personality traits by paced breathing and su-soku practice: A three-way crossover study. Compl Ther Med. 2021;63:102778. 10.1016/j.ctim.2021.102778.

26. Posse S, Olthoff U, Weckesser M, Jäncke L, Müller-Gärtner HW, Dager SR. Regional dynamic signal changes during controlled hyperventilation assessed with blood oxygen level-dependent functional MR imaging. AJNR Amer J of Neuroradiol. 1997;18(9):1763–1770.

27. Rotenstein LS, Torre M, Ramos MA, et al. Prevalence of Burnout Among Physicians: A Systematic Review. JAMA. 2018;320(11):1131–1150. doi:10.1001/jama.2018.12777

28. Russo MA, Santarelli DM, O’Rourke D. The physiological effects of slow breathing in the healthy human. Breathe (Sheff). 2017;13(4):298–309. doi:10.1183/20734735.009817

29. Shaffer F, Ginsberg JP. An Overview of Heart Rate Variability Metrics and Norms. Front Public Health. 2017;5:258. doi:10.3389/fpubh.2017.00258

30. Shanafelt TD, Dyrbye LN, West CP. Addressing Physician Burnout: The Way Forward. JAMA. 2017;317(9):901–902. doi:10.1001/jama.2017.0076

31. Teresi Ja, Yu X, Stewart AL, Hays RD. Guidelines for Designing and Evaluating Feasibility Pilot Studies. Med Care. 2022;60(1):95–103. doi:10.1097/MLR.0000000000001664.

32. Thabane, L., Lancaster, G. A guide to the reporting of protocols of pilot and feasibility trials. Pilot Feasibility Stud. 2019;37:5. doi10.1186/s40814-019-0423-8

33. Waxenbaum JA, Reddy V, Varacallo MA. Anatomy, Autonomic Nervous System. StatPearls. July 24, 2023. Accessed 11 1, 2025. https://www.ncbi.nlm.nih.gov/books/NBK539845/

34. Weiner, B.J., Lewis, C.C., Stanick, C. et al. Psychometric assessment of three newly developed implementation outcome measures. Implementation Sci. 2017;108:12. doi10.1186/s13012-017-0635-3

35. Woo T, Ho R, Tang A, Tam W. Global prevalence of burnout symptoms among nurses: A systematic review and meta-analysis. J Psychiatr Res. 2020;123:9–20. doi:10.1016/j.jpsychires.2019.12.015

36. Yildiz, S., Grinstead, J., Hildebrand, A. et al. Immediate impact of yogic breathing on pulsatile cerebrospinal fluid dynamics. Sci Rep. 2022;12(1):10894 10.1038/s41598-022-15034-8

37. Yoo SS, Gujar N, Hu P, Jolesz FA, Walker MP. The human emotional brain without sleep--a prefrontal amygdala disconnect. Curr Biol. 2007;17(20):R877–R878. doi:10.1016/j.cub.2007.08.007

38. Zaccaro A, Piarulli A, Laurino M, et al. How Breath-Control Can Change Your Life: A Systematic Review on Psycho-Physiological Correlates of Slow Breathing. Front Hum Neurosci. 2018;12:353. doi:10.3389/fnhum.2018.00353

